# A Quantitative Study of Factors Influencing Myasthenia Gravis Telehealth Examination Score

**DOI:** 10.1101/2024.07.24.24310934

**Authors:** Marc Garbey, Quentin Lesport, Helen Girma, Gülşen Öztosun, Henry J. Kaminski

## Abstract

**Introduction:** The increasing utilization of telemedicine offers an opportunity of video-recording to then perform post-hoc quantitative analysis of all aspects of the patient-examiner interaction. Presently, telemedicine examinations are adapted from the standard clinical visit, but their benefits and deficiencies have not been rigorously assessed.

**Methods:** We utilized a bank of 54 subjects with myasthenia gravis (MG) each having two telemedicine examinations by neuromuscular experts with recording of the MG Core Examination and the MG Activities of Daily Living. We then applied a broad spectrum of artificial intelligence algorithms from computer vision and speech analysis to natural language processing to generate quantitative metrics of the digital records.

**Results:** We successfully developed a technology to assess video examinations. Although the overall MG-CE examination scores were consistent across examiners, individual metrics showed significant variability, with up to a 25 percent variation in scoring within the MG-CE’s range. There was wide range of compliance with MG ADL instructions across examiners.

**Discussion:** We are able perform digital analysis of neuromuscular examinations and identified variations in outcome measure variations based on the examiner’s instructions, video recording limitations, and the severity of the patient’s disease. Particularly noteworthy is the high standard deviation in scores for examinations of patients with low disease severity.

---

Telemedicine and digital outcome metrics are being developed for many diseases,^1–3^ including myasthenia gravis (MG)^4,5^ with the promise to enhance performance of clinical trials and be adapted in routine clinical care. During the COVID-19 pandemic, many clinical trials were forced to rapidly move to telemedicine follow-up of subjects with concomitant uncertainty of ability to reliably execute clinical outcome metrics.^6^ Now, there is increasing acceptance of telemedicine with benefits of increased access, patient convenience, and closer follow-up of complex patients. An often-overlooked advantage of video visits, compared to in-person visits, is the recording of examinations to generate digital data sets, which are archivable for detailed analysis.

MG is particularly well-suited for the application of remote monitoring and need to improve clinical outcome measures.^7^ MG has highly variable disease manifestations and its severity may fluctuate over the course of a day and over weeks with the potential for severe exacerbations leading to hospitalizations and mortality. Clinical trials of MG are further challenged by its rarity leading to a need for a large number of centers recruiting with only a limited number of subjects per site. Further, there is a great burden to assure examiners, who lack experience in the disease, to accurately and consistently perform outcome measures. Variability in a clinical outcome measure translates to the need for greater subject number for statistical power to evaluate a primary outcome. The field of MG acknowledges that there are several sources of variability in outcome measure performance and a need to improve consistency.^8^

We took advantage of a unique data set of video recordings performed to assess the reproducibility of the MG core examination (MG-CE), which had been recommended for telemedicine evaluations.^9^ Using a broad spectrum of artificial intelligence algorithms related to computer vision, signal analysis, and natural language processing, we have constructed systems to quantitate aspects of the examination.^10,11^ These videos were originally obtained to assess the reproducibility of examination scores among the examiner who performed the telemedicine evaluation and then raters viewing the videos only (NCT05917184). Using these video recordings, we assessed sources of variations in assessments performed by the examiners. Here, we propose a digital technology to extract quantitative unbiased metrics that evaluate the limitations of the video technology, but more interestingly, the human examiner influence on the examination performance. Human factor studies rely on “expert” observations, but mitigating subjectivity by leveraging digital tools would be highly beneficial to yield unbiased, reproducible, quantitative data, and thereby enhance the objectivity of an examination.

## Methods

### Subjects and Video Recording

All subjects had been diagnosed with MG and confirmed by serum autoantibody elevations or electrophysiology. Each subject was asked to have stable internet connection and access to either a tablet or laptop with webcam. We used Zoom recordings (Zoom Video Communications, San Jose, CA) of subjects undergoing the MG-CE and MG-Activities of Daily Living Score (NCT05917184).^9^ which had been performed twice within 7 days except for one patient with 39 days between videos. For details of video recording analysis, please see our previous publications.^10,11^ The MG-CE evaluation assesses eight major domains, 2 ocular, 3 bulbar, 1 respiratory, and 2 limb and provides a severity scale of 0 (no deficit) to 3 (severe deficit) for each. Here, we added the score 4 do designate an examination could not be used for analysis. For both visits, each subject had been evaluated by the same neurologist with board certification in neuromuscular medicine and then recordings assessed by two reviewers and a third adjudicator if there were discrepancies in scoring. The complete dataset consisted of 102 videos of 51 different patients, performed by one of seven neurologists from five centers. All had been required to undergo training in performance of the MG-CE. All patients and examiners provided written consent and the study approved by the central institutional review board of MGNet at Duke University Institutional Review Board. We developed a Reproducibility Of Score (ROS) metric. Each individual score for one of the eight examinations was placed into one of two categories: 1) A reproductible score is defined as the examiner score agrees with the adjudicator, or agrees with both independent raters in the absence of an adjudicator score. 2) All other examinations are defined as non-reproductible. The classification provides a binary evaluation with either perfect agreement or not. To further refine our evaluation, we compute a dispersion of score defined as the number of different scores obtained by all three to four examiners for each test. We implicitly assume that coherent scores are better evaluations than those that are not. Dispersion of score among all graders is also a meaningful metric since we cannot assume that the adjudicator provides ground truth.

### Digital Algorithm of the Examination

AI processing of digital data was applied to the video image and the sound track using a broad spectrum of computer vision to analyze the video stream.^10,11^ In this investigation, we used a second voice signal analysis as well as Natural Language Processing (NLP) for the sound track using an AI transcription (AssemblyAI, San Francisco CA). The software provided a transcript that is annotated by time interval of each word during speech, an index of confidence for each word that has been reported in the transcript, and a classification of who is speaking. We used these tools to obtain metrics, such as the number of pixels in the Region Of Interest (ROI) for a patient examination, for example the eye lids for ptosis, if view of the ROI is complete or obscured, precise timing of the workflow, words used by the examiner, and adherence to the protocol. Data mining tools were used to assess the dispersion of the findings and identify interesting singularities.

### Assessment Related to the Influence of the Human Factor of the Examination Score

Three methodological steps were followed during the analysis. 1) We analyzed each examination to assess variability of the reproducibility of score and diversity in patient deficits for the data set. If the population model has too many of the patients exhibiting the same score value, for example 0 which indicates no abnormality, we cannot judge the potential impact of the human factor on the score. Similarly, if the reproducibility of the score is close to perfect for any category of the examination, there is no reason to look for a specific human factor that would affect the score.

We hypothesized that examiners would differ in consistency in scoring. We computed the overall score reproducibility for each examiner as measured by 1) adherence to protocol, 2) use of vocabulary, and 3) the workflow timing among across examinations. This evaluation is limited due to the small number of examiners.

We evaluated the individual examinations of the MG-CE to assess reasons for variations among the examiners, which could have related to inadequate data acquisition, such as 1) poor pixel resolution, 2) incorrect positioning of the patient in relationship to the camera, 3) poor adherence to instructions in performance of MG-CE.

## Results

### Score Distribution

Figure 1 displays the distribution of scores for each subject and exam category at each visit. With the exception of dysarthria, the population has a distributed representation of scores. Of 54 subjects, only four had confirmed signs of dysarthria, and five had suspicion, i.e. one positive score from either the observer or one reviewer without any confirmation by the adjudicator. Hence, we discarded the count-to-50 test from evaluation. We had previously concluded that the count-to-50 test is poorly reproducible and of limited clinical value.^12^

**Figure 1.**
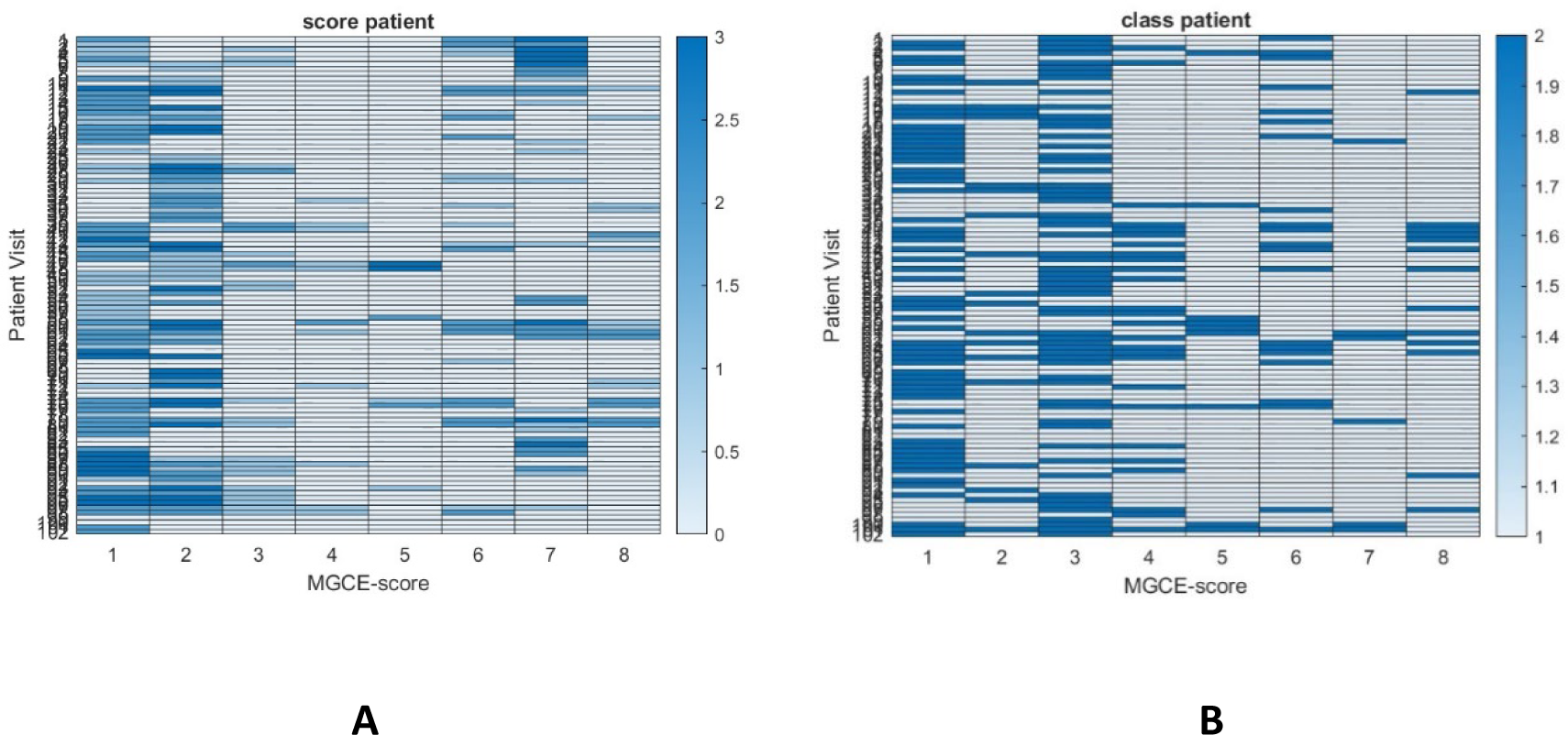
**A** Heat map of individual MG-CE scores by examiners of all 102 evaluations. Columns 1 to 8 represent the components of the MG-CE assessments: 1) Ptosis, 2) Diplopia, 3) Cheek Puff, 4) Tongue-to-Cheek, 5) Count-to-50, 6) Arm Extension, 7) Single-Breath-Count, and 8) Sit to Stand. **B** Reproducibility of the MG-CE scores. Reproducibility of the scores is evaluated in the context of the percentage of patients demonstrating abnormalities of each assessment and the distribution of scores. If the score of a test for the majority of patients is the same, the human factor influence on this test assessment cannot be established.

The average severity of the disease in each category provides context to the ROS. The population of patients had relatively low severity of disease. The tongue-to-cheek examination (column 4) and sit-to-stand exercise (column 8) make the classification of score reproducibility into Reproducible or Non-Reproducible less confident than high average severity, such as ptosis (column 1), diplopia (column 2) and single breath count (column 7).

We have simplified the characterization of potential human factor impact using a simple binary classification.^13,14^ For some examinations, we could have a ROS classification of four as examiner scores varied from 0 to 3.

### Holistic Attributes of the Examiner

We assessed the relationship of the score reproducibility with metrics of the examiner performance (Figure 2). There were seven examiners. Figure 3A shows the overall performance of each examiner based on their ROS score across all tests with variation from 0.6 to 0.8 on average, which is 20% to 27% of the range of the scoring. In Figure 3 the red bar corresponds to examiners with more than 25 years and black bar corresponds to examiners with less than 12 years since completion of fellowship training. Experience had little effect on score reproducibility. The most junior examiner was within the average range. The number of evaluations performed by examiners ranged from four to 32 with a median of 16.

**Figure 2.**
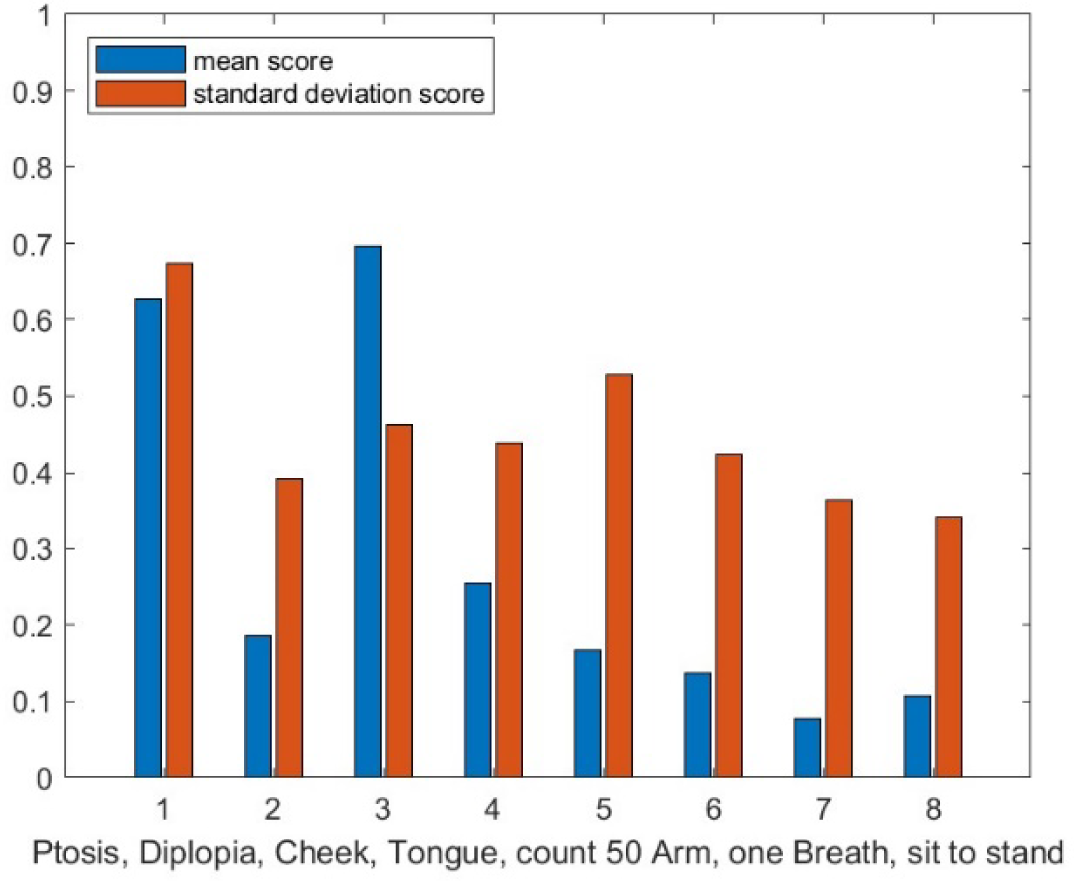
The variation in results among all evaluators is significant for each assessment. The mean and standard deviation of the score for all evaluations of all examiner. If score differences were uniformly distributed in the population, an average difference score of one for instance, would imply that there is always at least one grader who disagrees among all graders. Conversely, when the standard deviation is large and the mean score is low, disagreement on scores for certain patients can be notably pronounced.

**Figure 3.**
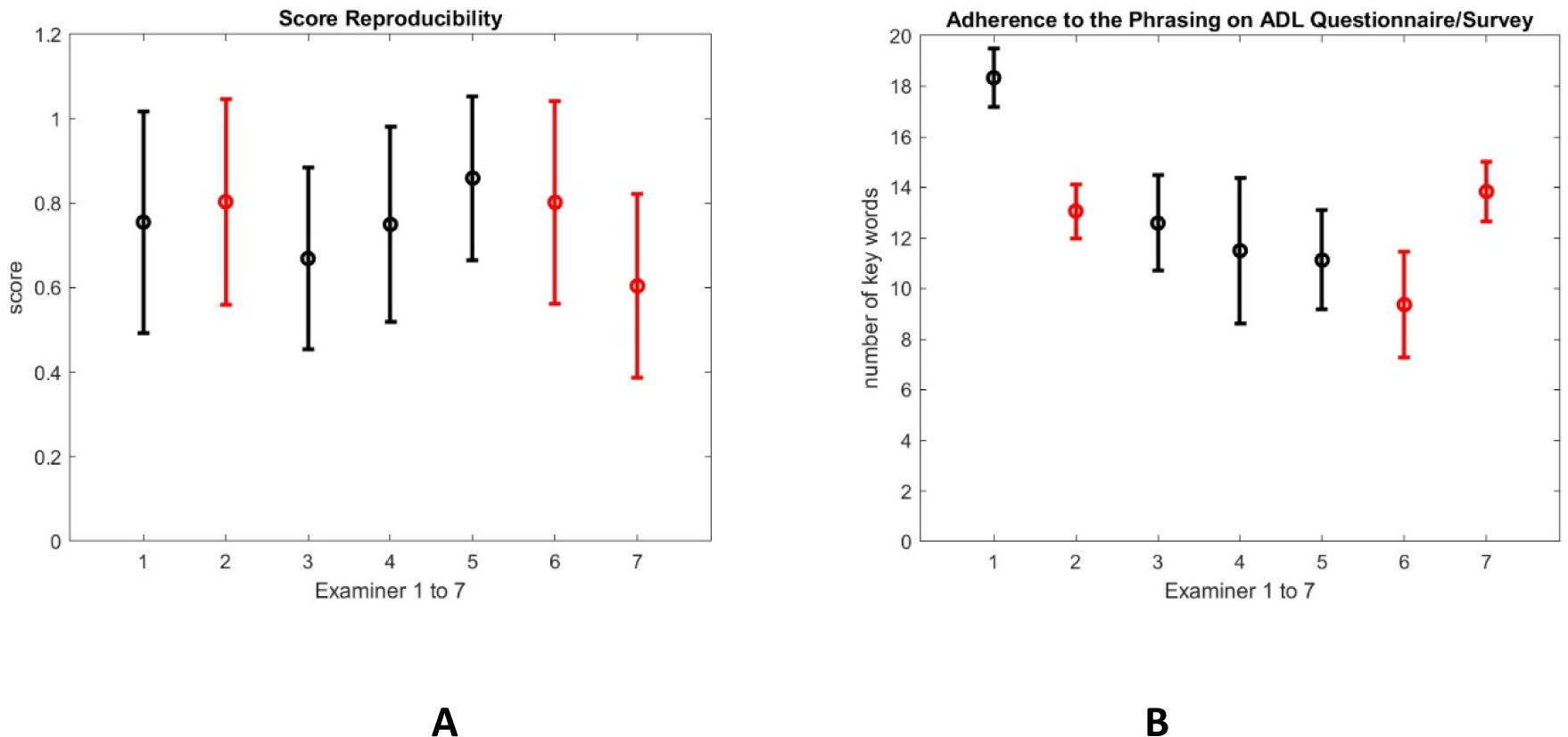
Assessment of MG-ADL Performance. **A)** Average reproducibility score per examiner and standard deviation among each of the assessments. **B)** Examiner adherence to the protocol based on the number of 20 pre-determined keywords they used while administering 8 questions of the Activities of Daily Living (ADL) questionnaire. Adherence to protocol is determined by the occurrence of the key words which are a part of the ADL and are detected through transcript of the examiner’s voice. Red bars correspond to examiners with greater than 25 years since completion of fellowship training, whereas the black bars correspond to examiners with less than 12 years.

### Assessment of MG-ADL Evaluation

NLP allowed for assessment for adherence to the MG-ADL instructions. This measure varied from 0 to 20, with 20 indicating the best adherence. This measure was based on the following key words of the MG-ADL questionnaire that should be placed in transcript in the correct order and time window: (Q1) slurred nasal speech, (Q2) fatigue jaw chewing, (Q3) choking swallow, (Q4) shortness breath, (Q5) brushing teeth, (Q6) trouble chair toilet, (Q7) double vision, (Q8) and drooping or droopy eyelids.^15,16^ Figure 3B indicates adherence to protocol was not associated with years of experience since completion of training or ROS performance.

We tested the possibility that the ROS performance might be related to the total time of the examination (Figure 4). Indeed, this elapsed time varied greatly across examinations. This may relate to severity of fatigue across subjects and the subject requiring recovery time between examinations. The second MG-CE exam was in average 20% faster than the first.

**Figure 4.**
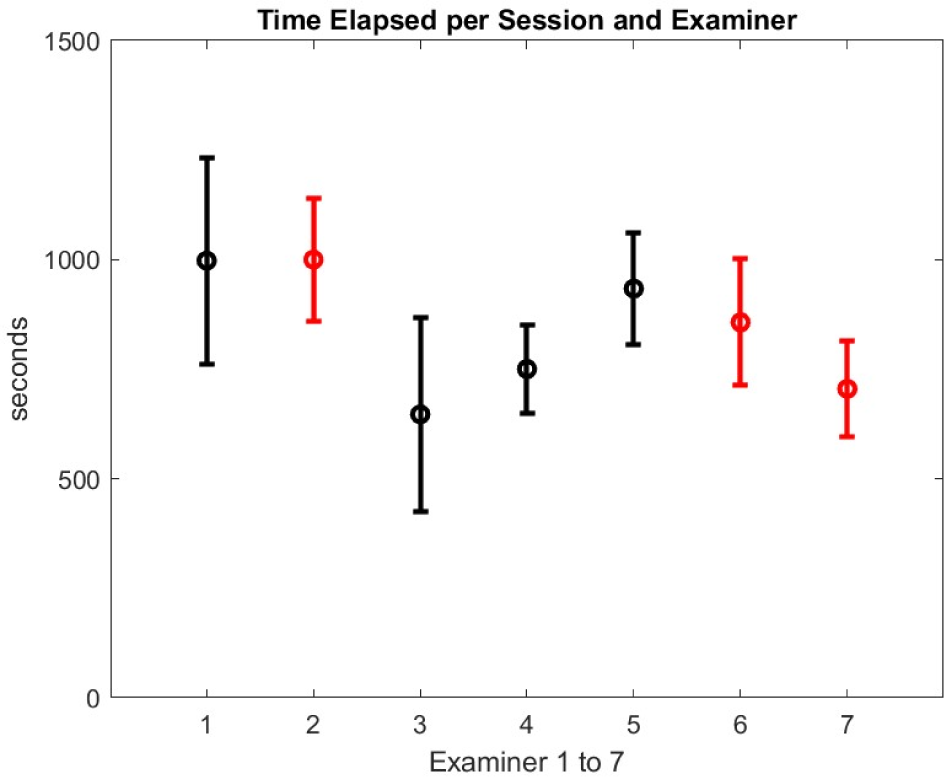
Average and standard deviation of the time elapsed in seconds per telemedicine session for each examiner. Red bars correspond to examiners greater than 25 years since completion of fellowship training, black bars those with less than 12 years.

We evaluated the fuzziness of the language used by the examiner doctor as defined by Icahn and colleagues^17^ and the sentiment metric ^18^ of the examiner speech but could not find any global patterns that would relate the ROS performance. The apparent variety of behavior makes essentially every doctor unique. We will next concentrate our analysis on specific examination in order to reduce the complexity of the problem and draw some useful conclusion that are independent of the doctor.

### Specific Exams of MG CE

The results of ocular and limb examinations allowed us to perform a more detailed analysis.

The number of pixels in the ROI varied greatly across videos depending on the distance between the patient and the camera. Figure 5 shows log normal distribution of the width of the patient eyes expressed in pixels. To investigate the ROS relationship with the video resolution, we split the data set into two categories about the same size: low pixel value that corresponds approximatively to the first two bars of the histogram of Figure 6 and high pixel value.

**Figure 5.**
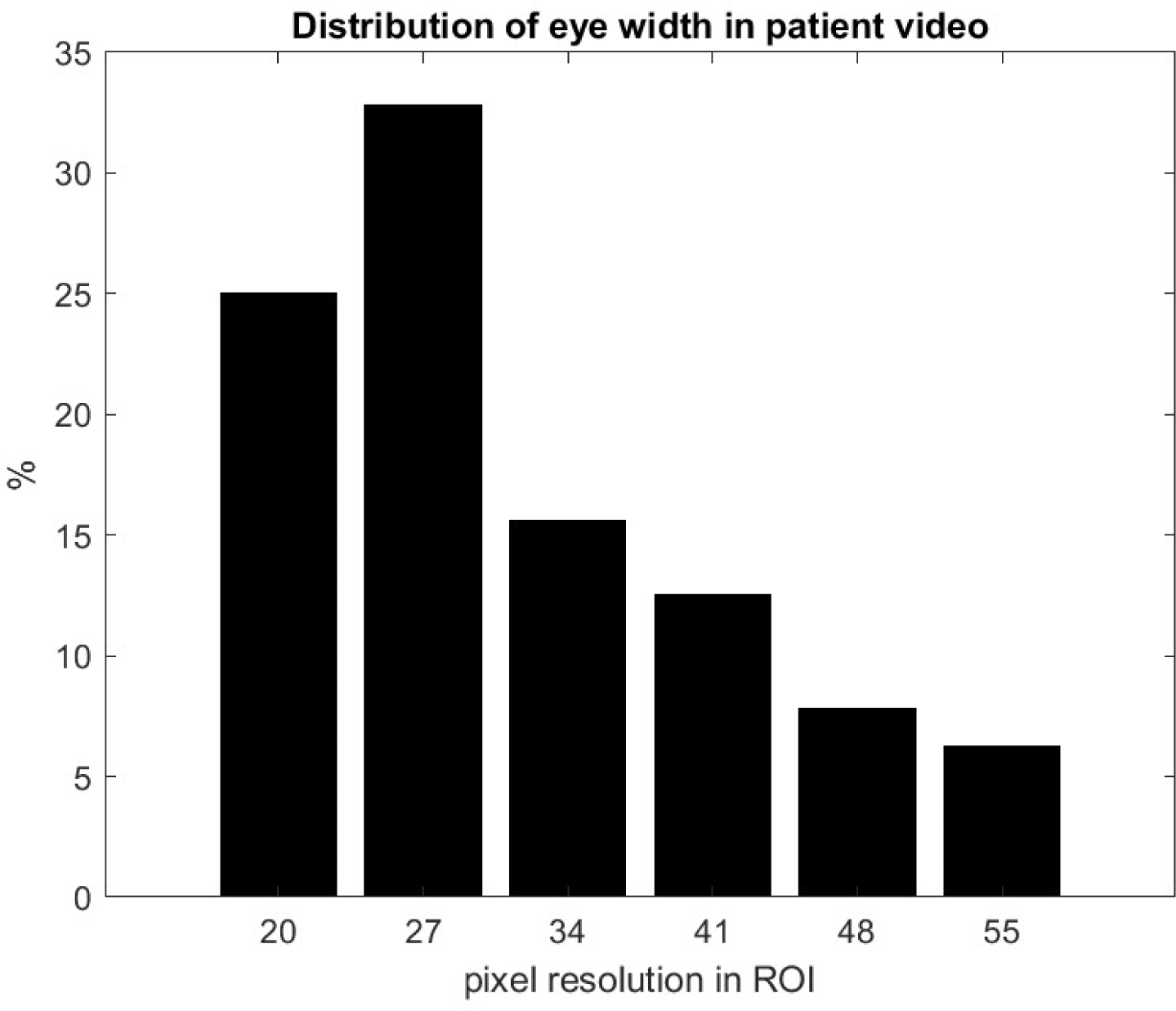
Resolution of region of interest (ROI) for the ptosis and diplopia assessments. The pixel resolutions are primarily affected by variations in the subjects’ distance from the camera and additional environmental factors, such as illumination.

**Figure 6.**
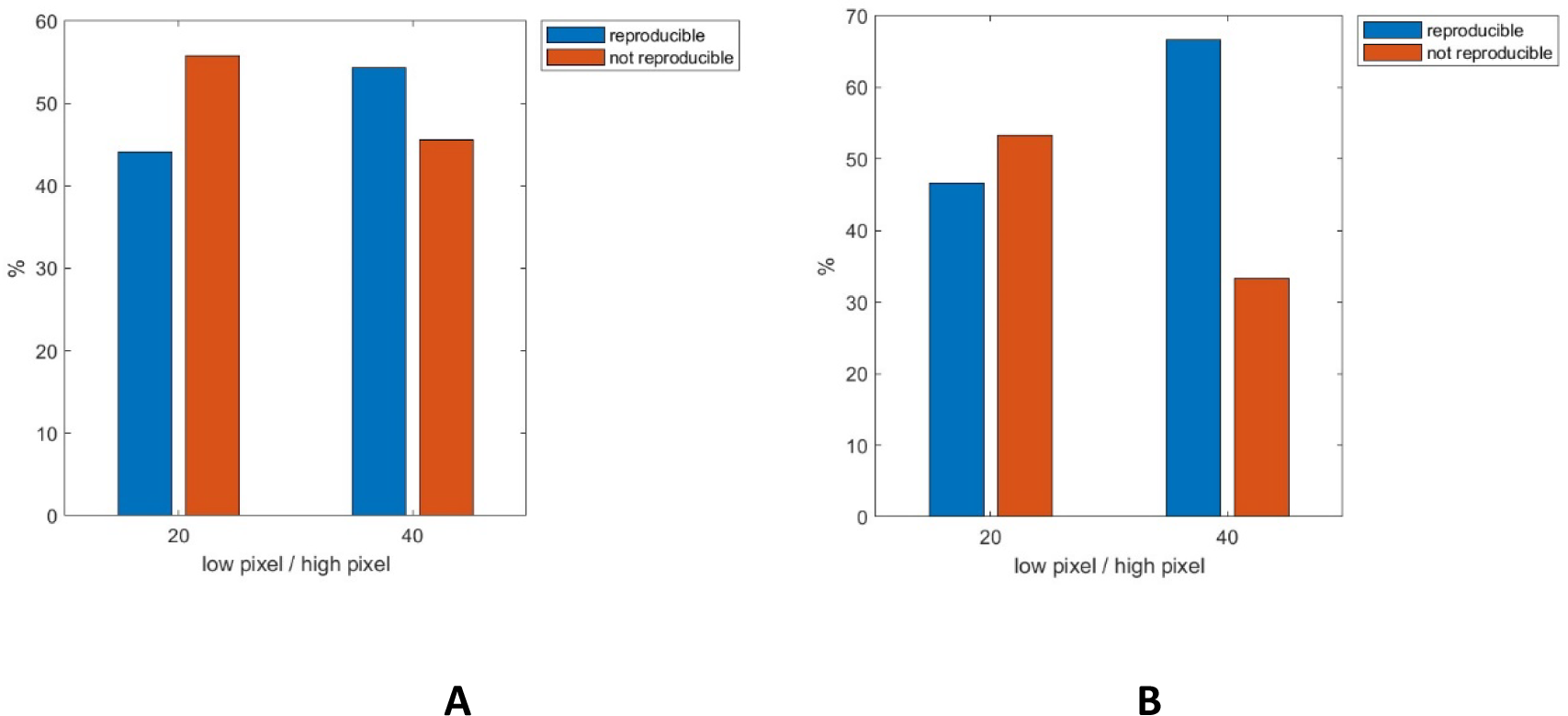
**A** Number of pixels in the region of interest (ROI) has a marginal effect on the score reproducibility for ptosis, although the percent reproducibility of that examination is low (42%). **B** Number of pixels in the ROI has a marginal effect on the score reproducibility for diplopia, although the percentage of reproducibility of that examination is high (83%). The observed high reproducibility percentage can be attributed to subjects reporting double vision themselves as opposed to identification of misalignment by the examiner. As a matter of fact, ocular misalignment is more difficult to judge in telemedicine as the subject tends to have greater lid drop when looking eccentrically.

Figure 6 A shows that the pixel resolution has limited impact on ROS with the Ptosis evaluation. These results agree with our previous work^10,11^ that shows that multiple factors impact the reproducibility of the ptosis score that turns out to have the largest uncertainty. We investigated the impact of the instructions of the examiner. It is essential that the subject to limit head movement during the 60 second test. We computed with NLP the frequency of key words related to the instructions – see Figure 2B. The more the examiner adhered to set instructions the better was the ROS, but this result is not robust. We speculate that the cause of poor ROS is multifactorial and many factors such as lighting, video resolution, subject anatomy contributes to the evaluation. To illustrate, we selected the group of patients for which there was a complete disagreement on the ptosis score from three evaluators. Of seven patients in that subset for which we had video archive, two had lower pixels values in the ROS, two did not look straight at the camera during the assessment, two struggled after the 60 seconds ptosis exercise to restore vision.

The Diplopia test is entirely dependent on patient report. Video observation by the examiner is unlikely to detect ocular misalignment. Improvement in pixel number show improvement in ROS (Figure 6B). As discussed in our previous publication,^11^ there is not necessarily a coincidence between ocular misalignment and the complaint of double vision.

For the arm extension, two separate metrics can be computed with digital video processing: how the arms drift downward and how long the subject can hold their arms extended during the two-minute test. The MG-CE score is reported as detection of drift at a certain time. Most videos did have a full view of the subject’s arms. Figure 7A shows that partial view of arm impacts the ROS. The reason likely lies with drift being more evident if the entire arm can be seen. Similarly, Figure 7B shows that for those patients who can hold the arms up to two minutes the ROS is less than for the ones who cannot. This suggests that the drift is more difficult to assess than the time of dropping the arms which is the major contribution to scoring.

**Figure 7.**
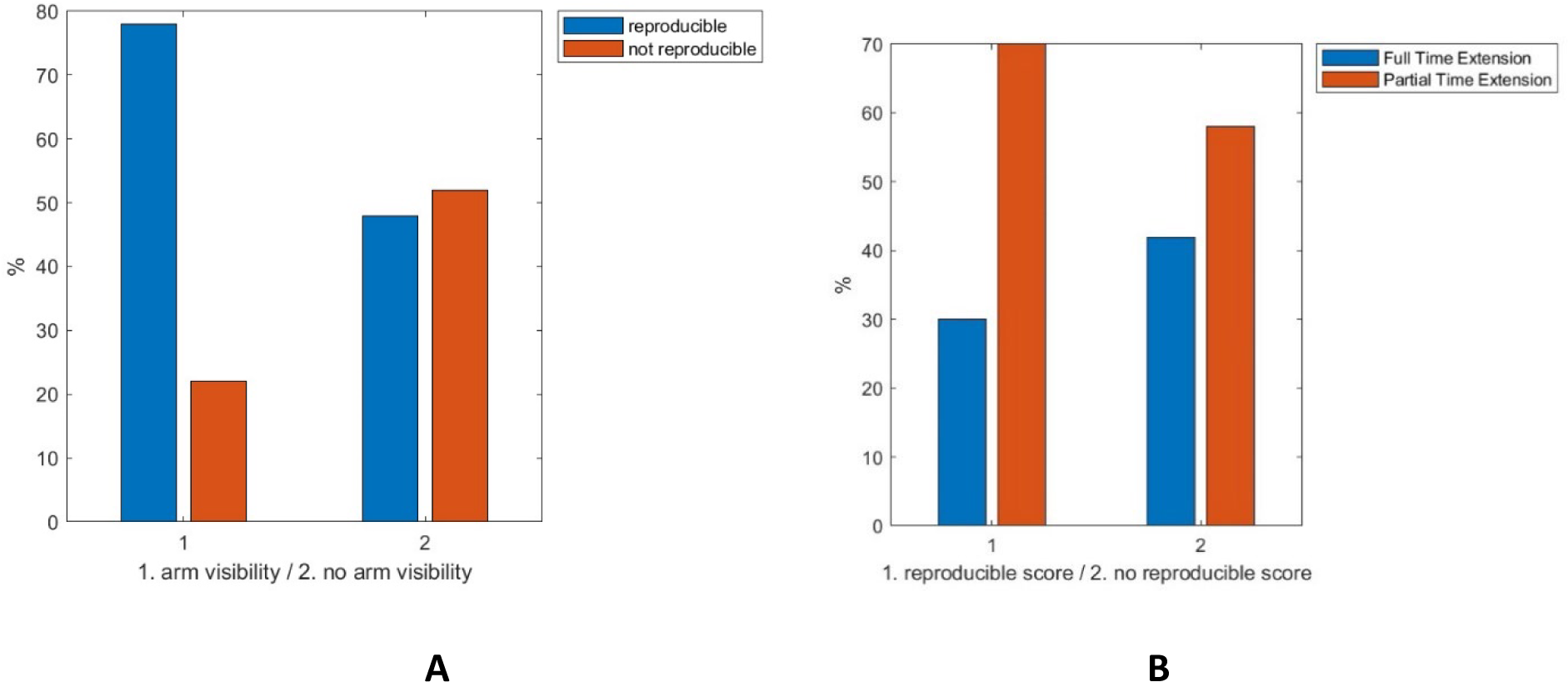
**A** Reproducibility of scores for the arm extension. The uncertainty of the score is attributed to the lack of visibility of the full length of extension. When the arms are not completely visible, there is a 50% percent chance that the score has no reproducibility. **B** Extension time impact on reproducibility of the score. When the arm extension assessment lasts for the full 120 seconds, assessing arm drift becomes challenging compared to when the duration of the extension lasts shorter. This difficulty arises because the evaluation protocol requires combining two distinct metrics: arm drift and duration of arm extension. In longer extensions, maintaining focus on both metrics simultaneously becomes more demanding, potentially leading to an obstacle in accurately assessing arm drift.

Evaluating the sit-to-stand of the patient seems straightforward and has indeed a high ROS (Figure 3). A limited view of the subject’s likely contributes to a poor ROS. For example, out of the ten subjects with both video’s visit with a full view of their bodies, only one had a scoring in the no ROS class.

The Cheek Puff test has a poor ROS (Figure 3) and appears to be of limited clinical value as assessed by telemedicine. If the patient does not close their lips and air leaks during the test it is difficult to detect. Further, there are no specific instruction for the duration of the test.

For the tongue in cheek test results are better, but the standard variation is fairly large compared to the mean of ROS. Because of varied anatomy as reflected in the mouth deformation, as determined in our previous analysis, this test cannot be quantified.

The speech tests have also been studied in our previous publication.^10^ Only roughly 10% of the patients are scored with dysarthria with this test. Our population sample has very low dysarthria severity distribution and is too small to make any conclusions.

The breath count lacks controls on frequency for counting which, makes it a poor evaluation of respiration. There is great variation across subjects in their ability to count loud and slow. The reproducibility of the score for one count breath should be perfect, since it is just a matter of writing how far the patient is counting. However, it was difficult for our algorithm detect the final number counted because of the rapid drop of the speech loudness. This is reflected in the transcript of the subjects counting as they become more out of breath.

## Discussion

Our study successfully developed a technology to analyze telemedicine-performed examinations, identifying outcome measure variations based on the examiner’s instructions, video recording limitations, and the severity of the patient’s disease. Intriguingly, although the overall MG-CE examination scores were consistent across examiners, individual metrics showed significant variability, with up to a 25 percent variation in scoring within the MG-CE’s range. Particularly noteworthy is the high standard deviation in scores for examinations of patients with low disease severity. This variability could substantially limit the ability to discern true treatment responses in clinical trials, especially for those with milder symptoms. Clinical trials for MG already face challenges in enrolling subjects with stable, moderate levels of weakness to avoid the need for rescue therapies or hospitalization, resulting in a skew towards lower severity cases.^7,8,19^ Our findings suggest that detecting improvement in these less severely affected subjects will be challenging. Furthermore, we found that established clinical outcome metrics for MG, including the MG-ADL, which has been the primary outcome measure in phase 3 trials, which led to FDA-approval of drugs for MG^20–22^ exhibit low discriminatory value for patients at the lower end of the severity spectrum.

Independent of the job title of the examiner (e.g. research coordinator, physician, subjects themselves) subjectivity is inevitable. Human factor studies are often overseen by external specialists who, while not always medical experts, employ a meticulous methodology to scrutinize the process.^23^ Unfortunately, human subjectivity in these studies with in person observation have their limitations. The present study provides an inclusive data set to address subjectivity. We took advantage of this unique video data set of examiners had *not* been the subject of the initial study REFERENCE (NCT05917184) allowing for a more real-world assessment. We were able to assess behaviors of examiners and identified variation in administration of the exam despite provision of a manual of administration and viewing of a training video. We did identify a training effect, in that all second examinations were typically 20% faster than the first. Reproducibility score was also not related to the length of examination time. Neither reproducibility nor adherence to the examination protocol, as assessed by natural language processing, was associated with years in training. We were not able to identify examiner vocabulary or sentiment associated with reproducibility of scoring. It is critical to appreciate that all our examiners are experts in neuromuscular disease, who would not necessarily need to behave in an identical manner to administer a familiar test. However, for novices and non-physicians, it is likely non-adherence to protocols could introduce even greater variation in outcome measures. To our knowledge, this is the first study to attempt to assess the impact of human examiner behavior on the examination of patients with a neurological disease.

Utilizing our previous algorithmic developments in digitalizing the MG-CE score, we have been able to quantify how factors such as pixel resolution limits, inadequate framing of patient views, poor sound acquisition, and ambiguous scoring definitions affect the reproducibility of the examination score. For instance, enhancing the pixel resolution during ocular motility assessments, such as ensuring subject proximity to the camera, could potentially increase the reproducibility of scoring by 15% for ptosis and 22% for diplopia. The benefit is even more pronounced for the arm extension test, where the success rate increases by 30% if the examiner assures that both arm of the patient are captured by the video frame. Overall, at the early stage of the development of our approach, those improvements are relatively modest, however they offer tangible areas to act upon. Our thorough examination of digital health data and extensive mining of the score database have uncovered a significant finding that aspects of the MG-CE protocol, specifically the Cheek Puff and Count-to-50 appear not to be effective measures in the telemedicine environment and by extension may not be even in the in-person clinic visit. Hence, it is imperative to consider substituting these assessments with more suitable alternatives, including video measurement of lip position and movements during speaking. However, ultimately despite a seemingly large data set of 102 videos, our results are compromised by the relatively small number of examiners performing the assessments. Also, the subjects overall had normal function on some tests, in particular the count-to-50 for evaluation of dysarthria, which limits our ability to assess the metric as an assessment of dysarthria. Further, our analysis is missing the non-verbal communication occurring between examiner and subject, as the examiner was not in the video feed to the subject during the examination. One could assume visual cues from the examiner could improve performance of some of the exam metrics.

We recommend that video archiving with appropriate security and privacy protections become part of subject examinations during clinical trials to support assessment of clinical trial outcome performance, but also identify quantitative evaluations. Such a process is not much different than archiving medical imaging. Recording with quantitation of telehealth provides a framework to enable clinical trials to more precisely compare assessments had potential to be profoundly impactful. Initially, such an approach could easily be applied to enhance training of study personnel prior to initiation of trials.

Ultimately, our primary goal is to refine our algorithms and establish a standardized protocol for collecting data from both patients/subjects and examiners during telemedicine examinations, and optimizing the time expenditure for subject and examiner. It is also crucial to ensure that software solutions and doctor-computer interfaces are non-intrusive. Underlying our work is that our assessments which are telemedicine based could be applied to international trials with trained examiners outside the local study site. As we define the optimal instructions, both by audio and visual instruction and by an AI-enabled avatar, performance of the MG-CE would become more consistent. Consistency would then reduce variability leading to less noise in an outcome measure, and if this were used as a primary outcome measure, then subject recruitment targets would be lower. Translated to clinical care, physician workload could be reduced to allow greater dedication of patient time to their immediate needs.

## Data Availability

All data produced in the present study are available upon reasonable request to the authors

## Acknowledgements

This work received partial support from the MGNet, a member of the Rare Disease Clinical Research Network Consortium (RDCRN), NIH U54 NS115054. Funding support for the DMCC is provided by the National Center for Advancing Translational Sciences (NCATS) and the National Institute of Neurological Disorders and Stroke (NINDS). Dr. Garbey was supported in part by NSF I-Corp award 838792 (47354/1/CCLS 91906F).

## Ethical Publication Statement

We confirm that we have read the Journal’s position on issues involved in ethical publication and affirm that this report is consistent with those guidelines.

